# Indoor and Outdoor Volatile Organic Compound Levels During and After the 2025 Los Angeles Wildfires

**DOI:** 10.1101/2025.03.31.25324857

**Authors:** Yuan Yao, Diane Garcia-Gonzales, Jing Li, Muchuan Niu, Michael Jerrett, Yifang Zhu

**Affiliations:** Department of Environmental Health Sciences, Jonathan and Karin Fielding School of Public Health, University of California, Los Angeles, California, USA

**Author notes:** Address correspondence to Michael Jerrett and Yifang Zhu.

## Abstract

The January 2025 Los Angeles urban wildfires caused extensive destruction and exposed millions to wildfire smoke containing hazardous volatile organic compounds (VOCs). To evaluate exposure risks, we conducted indoor and outdoor VOC sampling at 24 locations during three phases: active burning, smoldering, and off-gassing. Outdoor benzene concentrations peaked during active burning but remained below OEHHA health thresholds. In contrast, indoor BTEX concentrations increased during smoldering and remained elevated during the off-gassing phase, particularly in uninhabited homes inside burn zones, suggesting persistent emissions from smoke-impacted materials. These findings raise concerns about indoor air quality post-wildfire and the potential for prolonged exposure. We recommend ventilating homes and using HEPA and activated charcoal air purifiers before reoccupying fire-affected residences. Our results highlight the need for targeted mitigation and ongoing monitoring to protect public health during wildfire recovery.

## Introduction

The Los Angeles urban wildfires (LA Fires) that began on January 7, 2025, have now become collectively one of the most destructive disasters in U.S. history. The Palisades Fire burned over 23,707 acres in the Western part of the region, while the Eaton Fire in the Northeastern part of the region scorched approximately 14,021 acres.^1^ Together, these fires destroyed more than 16,000 structures, claimed at least 29 lives, and exerted untold costs to the health and wellbeing of millions of residents who experienced high levels of wildfire smoke exposure.

What made these urban wildfires particularly concerning was the potential toxicity of the resulting smoke, which likely contained volatile organic compounds (VOCs), such as benzene, toluene, ethylbenzene, and xylenes (BTEX), some of which are carcinogenic and harmful to human health.^2^ Uncertainty about the magnitude and distribution of VOCs during and after the fires has heightened community anxiety over short- and long-term health effects. Moreover, recent studies have shown that indoor VOCs levels can persist long after wildfires are contained, often exceeding outdoor concentrations.^3^ Some of these VOCs have been linked to physical symptoms lasting up to six months after residents return to homes near burn zones.^4^ Despite its importance, indoor air quality remains understudied in wildfire research, yet it is critical for informing public health mitigation strategies. To help fill this gap, we present field measurements of indoor and outdoor VOCs during and after the LA Fires.

## Methods

We conducted three rounds of one-week VOCs sampling using diffusion tubes to assess air quality changes during different phases: active burning (January 8–15, 2025), smoldering (January 24–31, 2025), and off-gassing (February 11–18, 2025). Paired indoor and outdoor samples were collected from 16 locations near the Palisades Fire and 9 locations near the Eaton Fire. VOC analysis was performed using thermal desorption with gas chromatography. Four samples near the Palisades Fire and three samples near the Eaton Fire were excluded from the data analysis due to being either below the detection limits or overloaded, resulting in a final sample set of 43 from 24 locations. Results were compared to regional background levels before the fires, including two locations sampled both pre- and during the fires.

## Results and Discussion

Figure 1 shows the spatial distribution of indoor and outdoor BTEX concentrations near both the Palisades and Eaton Fires. Concentrations of individual BTEX compounds are summarized in Table 1. Across the three study phases—active burning (Jan 8–15), smoldering (Jan 24–31), and off-gassing (Feb 11–18), BTEX concentrations exhibit distinct patterns indoors and outdoors.

**Table 1.** Indoor and Outdoor Concentrations of Benzene, Toluene, Ethylbenzene, and Xylenes (BTEX) During Different Phases of the LA Fires.

| Phase | Indoor or outdoor | Statistics | Benzene (ppb) | Toluene (ppb) | Ethylbenzene (ppb) | m,p-Xylene (ppb) | o-Xylene (ppb) | BTEX (ppb) |
| --- | --- | --- | --- | --- | --- | --- | --- | --- |
| 1 | Indoor | Mean ± SD | 0.28 ± 0.15 | 0.32 ± 0.21 | 0.02 ± 0.01 | 0.04 ± 0.03 | 0.01 ± 0.01 | 0.67 ± 0.40 |
|  |  | Range | 0.13 – 0.48 | 0.06 – 0.53 | 0.01 – 0.04 | 0.01 – 0.07 | 0.00 – 0.03 | 0.23 – 1.13 |
|  | Outdoor | Mean ± SD | 0.32 ± 0.13 | 0.29 ± 0.20 | 0.03 ± 0.02 | 0.06 ± 0.04 | 0.02 ± 0.01 | 0.71 ± 0.38 |
|  |  | Range | 0.15 – 0.48 | 0.08 – 0.55 | 0.01 – 0.06 | 0.02 – 0.12 | 0.00 – 0.04 | 0.25 – 1.19 |
| 2 | Indoor | Mean ± SD | 0.15 ± 0.05 | 0.56 ± 0.24 | 0.03 ± 0.03 | 0.15 ± 0.11 | 0.03 ± 0.03 | 0.91 ± 0.29 |
|  |  | Range | 0.10 – 0.23 | 0.25 – 0.90 | 0.00 – 0.07 | 0.03 – 0.27 | 0.00 – 0.07 | 0.49 – 1.38 |
|  | Outdoor | Mean ± SD | 0.15 ± 0.03 | 0.48 ± 0.28 | 0.02 ± 0.03 | 0.09 ± 0.11 | 0.01 ± 0.02 | 0.75 ± 0.38 |
|  |  | Range | 0.12 – 0.18 | 0.16 – 0.70 | 0.00 – 0.05 | 0.00 – 0.22 | 0.00 – 0.04 | 0.33 – 1.07 |
| 3 | Indoor | Mean ± SD | 0.08 ± 0.03 | 0.53 ± 0.36 | 0.03 ± 0.07 | 0.14 ± 0.26 | 0.03 ± 0.07 | 0.79 ± 0.48 |
|  |  | Range | 0.04 – 0.14 | 0.20 – 1.51 | 0.00 – 0.22 | 0.00 – 0.89 | 0.00 – 0.22 | 0.31 – 1.78 |
|  | Outdoor | Mean ± SD | 0.07 ± 0.02 | 0.24 ± 0.12 | 0.00 ± 0.00 | 0.01 ± 0.02 | 0.00 ± 0.00 | 0.32 ± 0.12 |
|  |  | Range | 0.05 – 0.10 | 0.05 – 0.39 | 0.00 – 0.00 | 0.00 – 0.06 | 0.00 – 0.00 | 0.11 – 0.51 |

**Figure 1.**
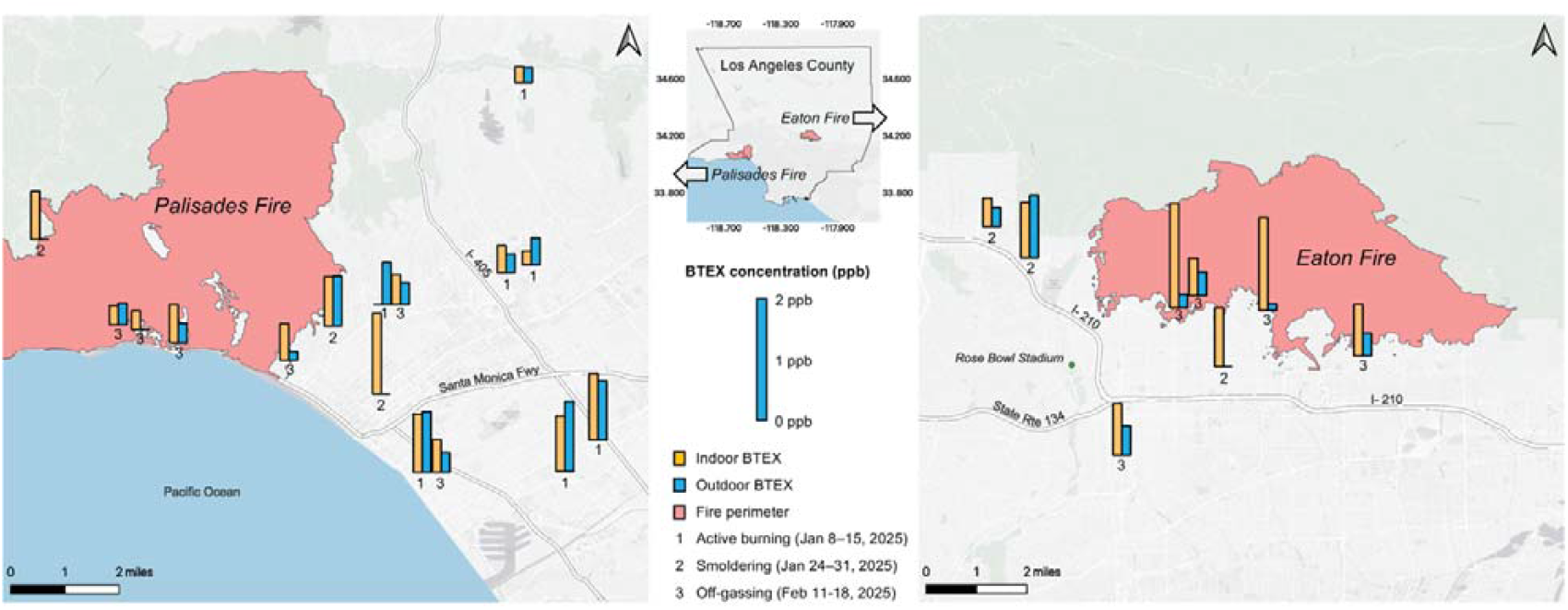
Indoor and Outdoor Concentrations of Benzene, Toluene, Ethylbenzene, and Xylenes (BTEX) near the Palisades and Eaton Fires. Data were collected during Phase 1 (January 8–15, 2025, active burning), Phase 2 (January 24–31, 2025, smoldering), and Phase 3 (February 11–18, 2025, off-gassing). Yellow bars represent indoor BTEX concentrations, while blue bars represent outdoor BTEX concentrations.

For outdoor concentrations, the average BTEX levels were 0.71, 0.75, and 0.32 ppb, during Phase 1, Phase 2, and Phase 3, respectively (Table 1). Notably, outdoor benzene levels during Phase 1 were 0.32 ppb exceeding the nearby Burbank Area level of 0.22 ppb and the Central LA level of 0.26 ppb, as reported in the MATES V study, conducted by the South Coast Air Quality Management District from May 2018 to April 2019.^5^ The area south of the Palisades burn zone experienced an increase in total BTEX compounds, with a 192% increase in benzene during the active burning period (January 8–15, 2025) compared to the pre-fire period (May 16–30, 2024). In contrast, the site east of the Palisades burn area showed only a slight 25% increase in benzene, likely due to dominant plume trajectory (see Figure 2 in Schollaert et al. 2025).^6^ Despite increased outdoor benzene concentrations observed, these levels remained below health standards set by the California Office of Environmental Health Hazard Assessment (OEHHA); however, the World Health Organization notes that because benzene is carcinogenic to humans, no safe threshold of exposure exists.^7^

In comparison, average indoor BTEX concentrations were 0.67, 0.91, and 0.79 ppb, during Phase 1, Phase 2, and Phase 3, respectively. During the active fire event, indoor BTEX levels were lower than outdoor levels, lending some support to public health advisories recommending that residents shelter indoors. During the smoldering phase, both indoor and outdoor BTEX concentrations increased, suggesting ongoing emissions. This pattern changed during the post-fire recovery phase, when uninhabited homes within the burn zones, particularly near the Eaton Fire, exhibited unexpectedly higher indoor BTEX concentrations compared to outdoor air, likely due to off-gassing from smoke-impacted materials. This finding highlights a potential health risk: while residents may feel safe returning home after wildfires have been extinguished, they could be exposed to lingering VOCs indoors. To reduce indoor exposure, before returning home residents should ventilate homes regularly by opening windows, running central air systems (heating, ventilation, and air conditioning—HVAC) equipped with filters rated Minimum Efficiency Reporting Value (MERV) 13 or higher, and using High-Efficiency Particulate Air (HEPA) purifiers when possible. These measures can help accelerate off-gassing and improve indoor air quality.

## Data Availability

All data produced in the present study are available upon reasonable request to the authors.

## Acknowledgments

Jonathan Fielding, Miriam E. Marlier, Beate Ritz, Feng Gao, Christina Batteate, Department of Environmental Health Sciences, Fielding School of Public Health, University of California Los Angeles, and the Center for Healthy Climate Solutions. Michael Kleeman, Thomas M. Young, Christopher P. Alaimo, University of California Davis. Jiachen Zhang, Department of Civil and Environmental Engineering, University of Southern California. This study was partially funded by the Spiegel Family Fund, the Gordon and Betty Moore Foundation, and the R&S Kayne Foundation.

